# ChatGPT achieves comparable accuracy to specialist physicians in predicting the efficacy of high-flow oxygen therapy

**DOI:** 10.1101/2023.10.12.23296773

**Authors:** Taotao Liu, Yaocong Duan, Yanchun Li, Yingying Hu, Lingling Su, Aiping Zhang

## Abstract

**Rationale:** The failure of high-flow nasal cannula (HFNC) oxygen therapy can necessitate endotracheal intubation in patients. Timely prediction of the endotracheal intubation risk due to HFNC failure is critical for avoiding delays in intubation, therefore potentially decreasing mortality.

**Objectives:** To investigate the accuracy of ChatGPT in predicting the risk of endotracheal intubation within 48 hours after HFNC therapy and compare it with the predictive accuracy of specialist and non-specialist physicians.

**Methods:** We conducted a prospective multicenter cohort study based on the data of 71 adult patients who received HFNC therapy. We recorded patient baseline data, the results of blood gas analysis, and physiological parameters after 6-hour HFNC therapy. For each patient, this information was used to create a 6-alternative-forced-choice natural language questionnaire that asked participants to predict the risk of 48-hour endotracheal intubation using graded options from 1 to 6, with higher scores indicating a higher risk. GPT-3.5, GPT-4.0, respiratory and critical care specialist physicians and non-specialist physicians completed the same 71 questionnaires respectively. We then determined the optimal diagnostic cutoff point for each of them, as well as 6-hour ROX index, using the Youden index and compared their predictive performance using receiver operating characteristic (ROC) analysis.

**Results:** The optimal diagnostic cut-off points for GPT-4.0 and specialist physicians were determined to be ≥4. The precision of GPT-4.0 was 76.1% [specificity=78.6% (95%CI=52.4-92.4%); sensitivity=75.4% (95%CI=62.9-84.8%)]. The precision of specialist physicians was 80.3% [specificity=71.4% (95%CI=45.4-88.3%); sensitivity=82.5% (95%CI=70.6-90.2%)]. The optimal diagnostic cut-off points for GPT-3.5 and non-specialist physicians were determined to be ≥5, with precisions of 73.2% and 64.8% respectively. The area under the ROC (AUROC) of GPT-4.0 was 0.821 (95%CI=0.698-0.943), which was greater than, but not significantly (p>0.05) different from the AUROCs of GPT-3.5 [0.775 (95%CI=0.652-0.898)] and specialist physicians [0.782 (95%CI=0.619-0.945)], while was significantly higher than that of non-specialist physicians [0.662 (95%CI=0.518-0.805), P=0.011]. Grouping the patients by GPT-4.0’s prediction value ≥4 (high-risk group) and ≤3 (low-risk group), the 28-day cumulative intubation rate (56.00% vs. 15.22%, P<0.001) and 28-day mortality (44.00% vs. 10.87%, P<0.001) of the high-risk group were significantly higher than those of the low-risk group.

**Conclusion:** GPT-4.0 achieves an accuracy level comparable to specialist physicians in predicting the 48-hour endotracheal intubation risk in patients after HFNC therapy, based on patient baseline data and 6-hour parameters of receiving HFNC therapy. Large-scale studies are needed to further inspect whether GPT-4.0 can provide reliable clinical decision support.

---

High-flow nasal cannula (HFNC) oxygen therapy, a method used to deliver a heated, humidified, and high-flow air-oxygen mixture to patients, has been shown to be effective in treating hypoxemia and is widely applied in clinical practice due to its convenience and comfort[1, 2]. Recent studies have further explored the indications of HFNC therapy[3-5]. However, sequential treatment failure with HFNC therapy can lead to the need for endotracheal intubation in patients. If endotracheal intubation is delayed, it can increase mortality[6]. Therefore, it is critical to predict in advance the risk of endotracheal intubation in patients due to HFNC failure. Although recent studies have shown that the ratio of SpO_2_/FiO_2_ to respiratory rate (the ROX index) can be used to predict the efficacy of HFNC therapy[7], its predictive accuracy is only moderate, and its diagnostic cut-off points lack standardized criteria[8, 9].

While artificial intelligence (AI) shows promise in supporting clinical decision-making, the complex algorithms and the high learning costs hinder physicians without programming experience from using AI-assisted decision-making. Recent advances in natural language processing (NLP) tools, such as ChatGPT, enable physicians to use AI in a natural language manner. This means they can focus on recording medical data in natural language format that serves as prompts for NLP tools rather than bothering with complex algorithms. The potential of using ChatGPT to support clinical judgement of endotracheal intubation after HFNC therapy remains unexplored.

This study hypothesizes that the prediction of GPT-4.0 on the risk of 48-hour endotracheal intubation in patients after HFNC therapy, based on patient baseline data and parameters of receiving 6-hour HFNC therapy, is at least as good as that of physicians who are not specialized in respiratory and critical care. To test this hypothesis, we developed a natural language questionnaire based on 71 prospectively included patients receiving HFNC oxygen therapy from multiple centers, and obtained the predictions of the 48-hour endotracheal intubation risk from specialist physicians, non-specialist physicians, GPT-3.5, and GPT-4.0 to compare their predictive accuracy. The same 71 questionnaires were completed by GPT-3.5, GPT-4.0, specialist physicians (three specialists completed 71 questionnaires by each independently completing one part), and non-specialist physicians (Three non-specialists completed 71 questionnaires in the same manner).

## Methods

Medical research ethics approval was obtained from each center (the First Affiliated Hospital of Henan University of Science and Technology 2021-0241; Jiangyan Hospital Affiliated to Nanjing University of Chinese Medicine 2021-016). Patients or their family members provided informed consent, and the study was registered as a clinical trial (ChiCTR2100053027). Full study protocol can be accessed from https://www.chictr.org.cn.

### Patients

This cohort study prospectively included 73 patients from two Grade-A tertiary care and teaching hospitals. After excluding 2 patients, 71 patients receiving HFNC oxygen therapy (Respircare HUMID BH) were finally included in the study.

#### Inclusion criteria

Patients over 18 years old; receiving HFNC oxygen therapy due to various clinical needs; with or without type 2 respiratory failure.

#### Exclusion criteria

Patients received tracheostomy; Patients who refuse intubation during 48-hour HFNC therapy; Patients who request to withdraw from the study; incomplete data collection; Patients who received intermittent non-invasive ventilation during 48-hour HFNC therapy; Patients who received prone position ventilation during 48-hour HFNC therapy.

### Study design

Recorded baseline data includes: age, gender, body mass index (BMI), mechanical ventilation history, comorbidities, main diagnosis.

Recorded the initial HFNC oxygen therapy and treatment parameters at 6 hours include: blood gas analysis results, respiratory rate, heart rate, pulse oximetry (SpO_2_), blood pressure, fraction of inspired oxygen (FiO_2_), and oxygen flow and Glasgow Coma Scale (GCS) score. The endpoint of HFNC oxygen therapy observation will be either: tracheal intubation or tracheotomy; patient death; or 48 hours of HFNC treatment.

Clinical outcomes were followed up, including 1) the primary clinical outcome: tracheal intubation within 48 hours, and 2) secondary outcomes: time to tracheal intubation, time to death, 28-day tracheal intubation rate, 28-day mortality rate, and length of stay (LOS) in hospital. The follow-up endpoint was either patient death, discharge, or 28-day hospitalization.

We recorded the above data of 71 patients into 71 natural language questionnaires that asked participants to predict the 48-hour endotracheal intubation risk in patients after HFNC therapy, based on options that were graded from 1 to 6: 1) extremely unlikely to undergo endotracheal intubation, 2) unlikely to undergo endotracheal intubation, 3) possible not to undergo endotracheal intubation, 4) possible to undergo endotracheal intubation, 5 likely to undergo endotracheal intubation, and 6) extremely likely to undergo endotracheal intubation. One forced choice was required. A template of questionnaire was shown in Box 1.

Both GPT-3.5 and GPT-4.0 were used to predict the 48-hour endotracheal intubation risk by prompting the questionnaire contents. Besides, three respiratory and critical care specialists aged 30 to 40 independently completed 23 to 24 questionnaires each, and a total of 71 questionnaires were completed. Three non-specialist physicians aged 30 to 40 independently completed 23 to 24 questionnaires each, and a total of 71 questionnaires were completed (see Table 2 and Figure 2).

**Table 1.**
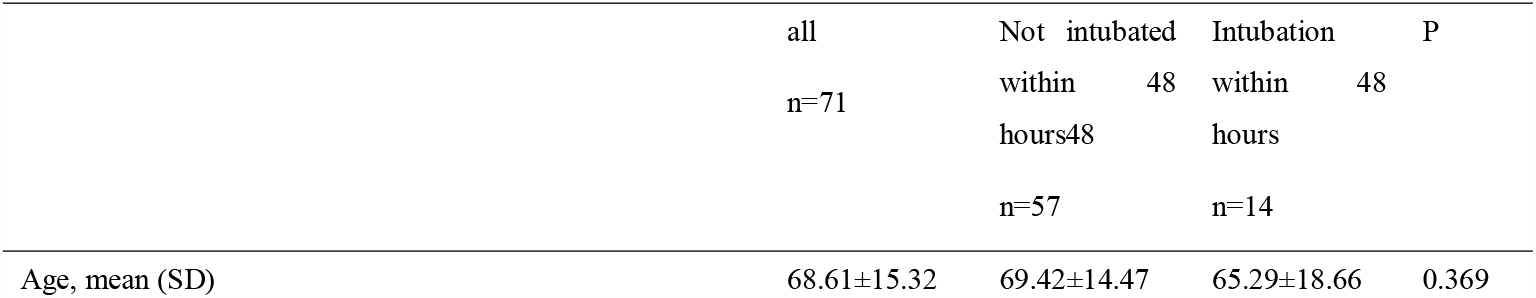
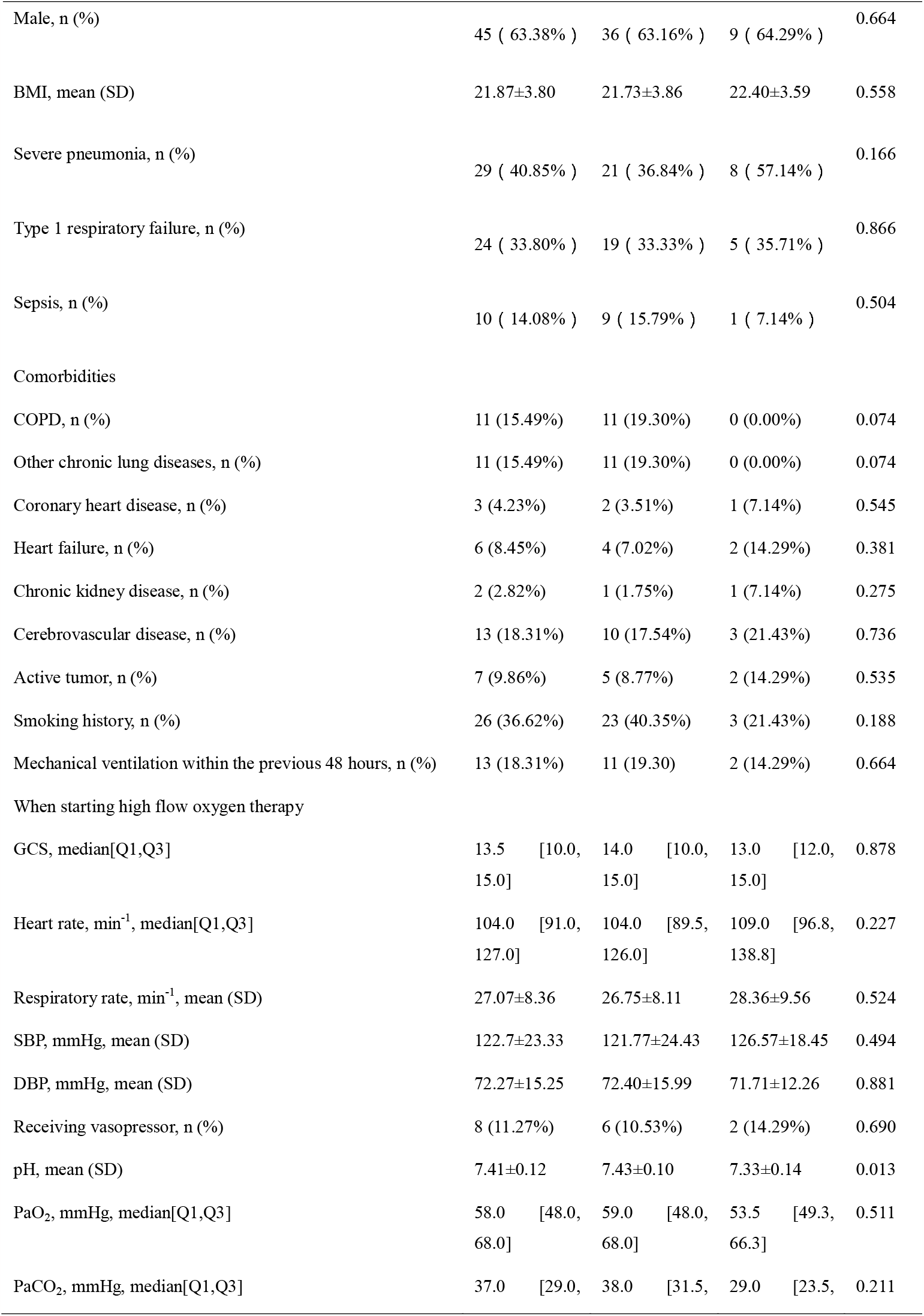
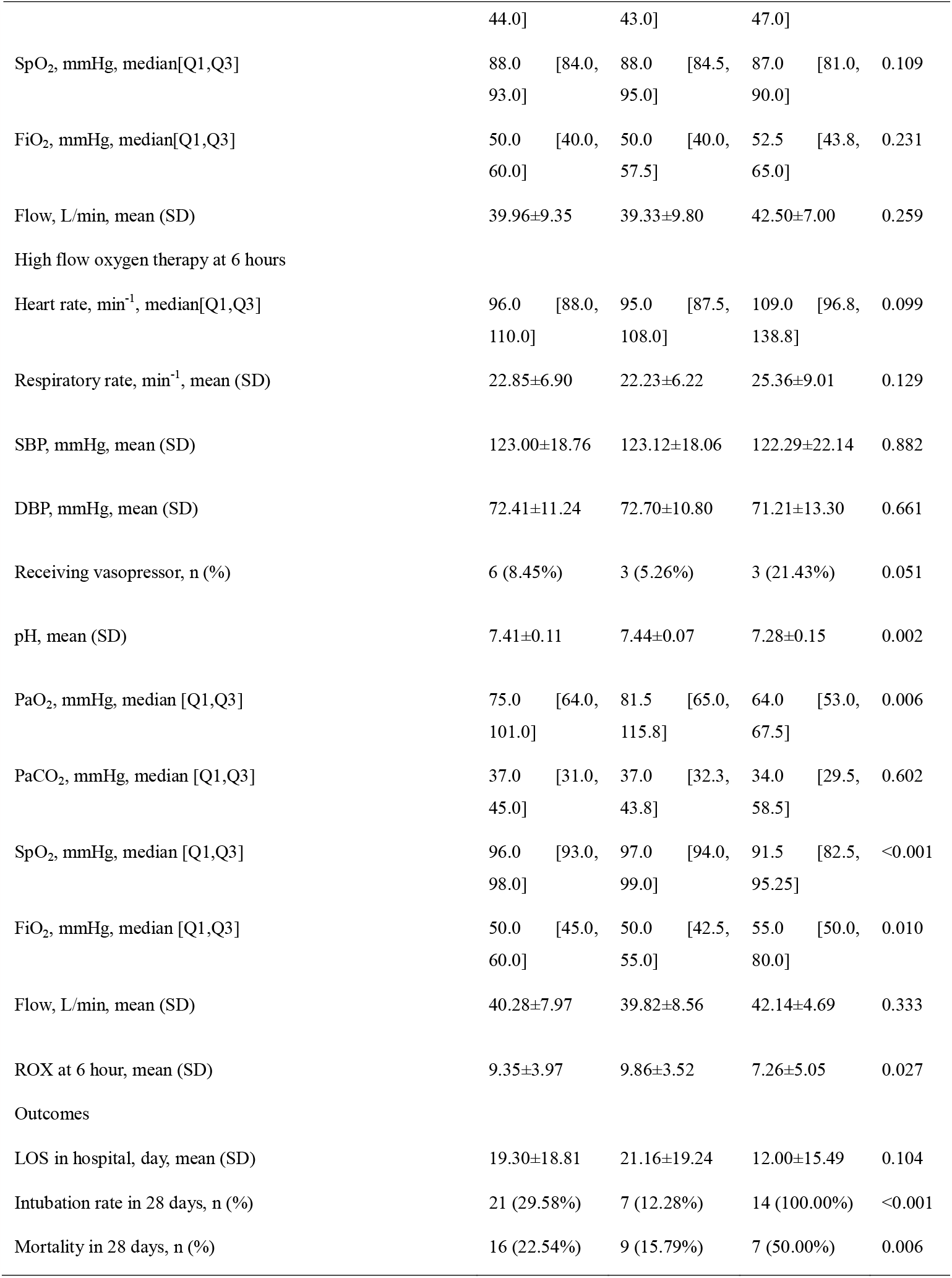
Clinical characteristics of patients with endotracheal intubation and without intubation within 48 hours of treatment.

**Table 2.** Prediction results.

|  | GPT 3.5's<br>prediction | 48-hour<br>intubation<br>in<br>practice | GPT 4's<br>prediction | 48-hour<br>intubation<br>in<br>practice | Specialist<br>physicians'<br>prediction | 48-hour<br>intubation<br>in<br>practice | Non-specialist<br>physicians'<br>prediction | 48-hour<br>intubation<br>in<br>practice |
| --- | --- | --- | --- | --- | --- | --- | --- | --- |
| 1 extremely<br>unlikely to<br>undergo<br>endotracheal<br>intubation | 0 | 0 | 0 | 0 | 13 | 2 | 14 | 0 |
| 2 unlikely to<br>undergo<br>endotracheal<br>intubation | 9 | 0 | 29 | 1 | 22 | 0 | 11 | 3 |
| 3 possible<br>not to<br>undergo<br>endotracheal<br>intubation | 3 | 0 | 17 | 2 | 16 | 2 | 10 | 1 |
| 4 possible to<br>undergo<br>endotracheal<br>intubation | 32 | 3 | 17 | 6 | 6 | 2 | 7 | 1 |
| 5 likely to<br>undergo<br>endotracheal<br>intubation | 27 | 11 | 7 | 4 | 6 | 3 | 14 | 5 |
| 6 extremely<br>likely to<br>undergo<br>endotracheal<br>intubation | 0 | 0 | 1 | 1 | 8 | 5 | 15 | 4 |

**Figure 1.**
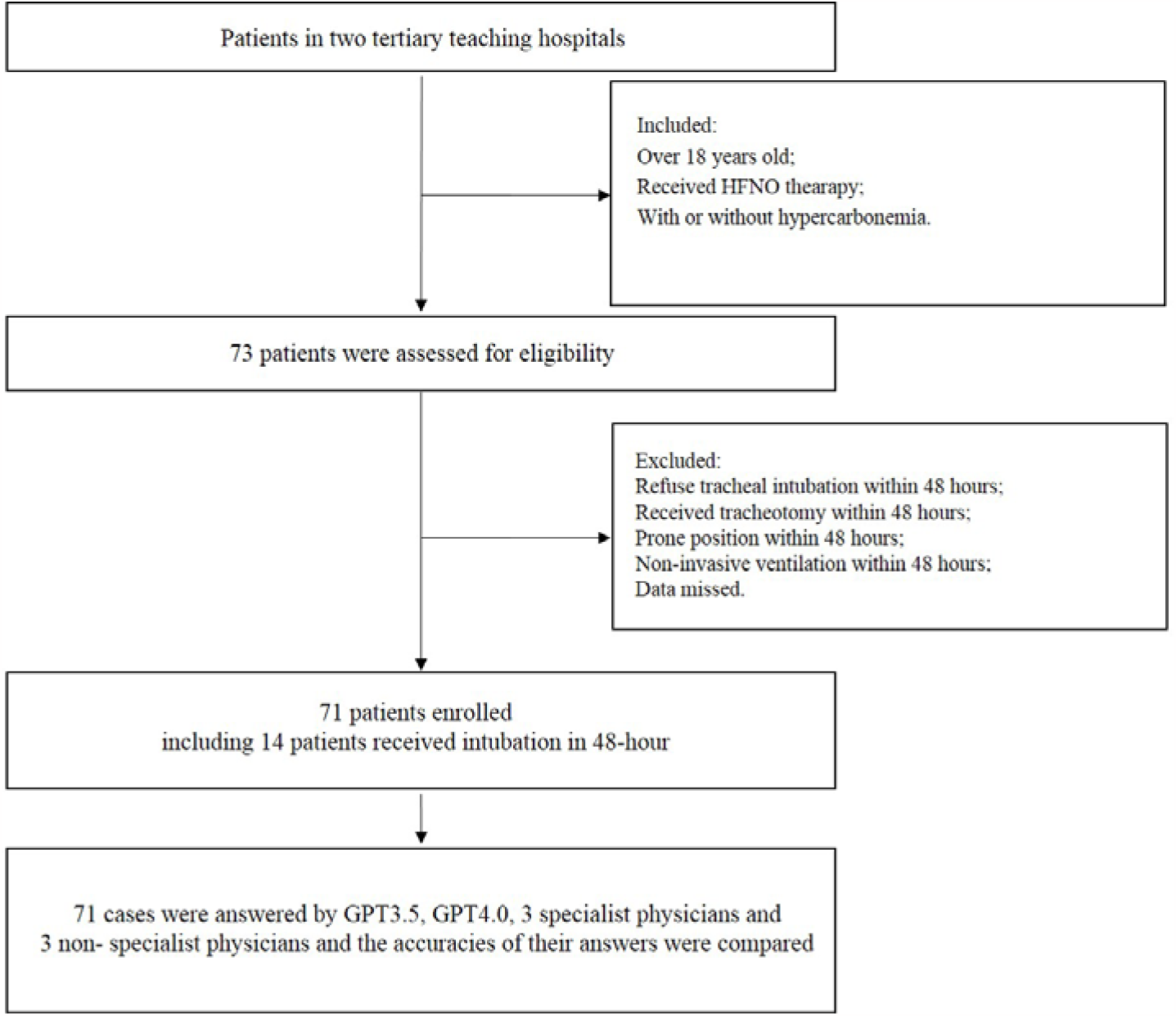
Flow chart of study

**Figure 2.**
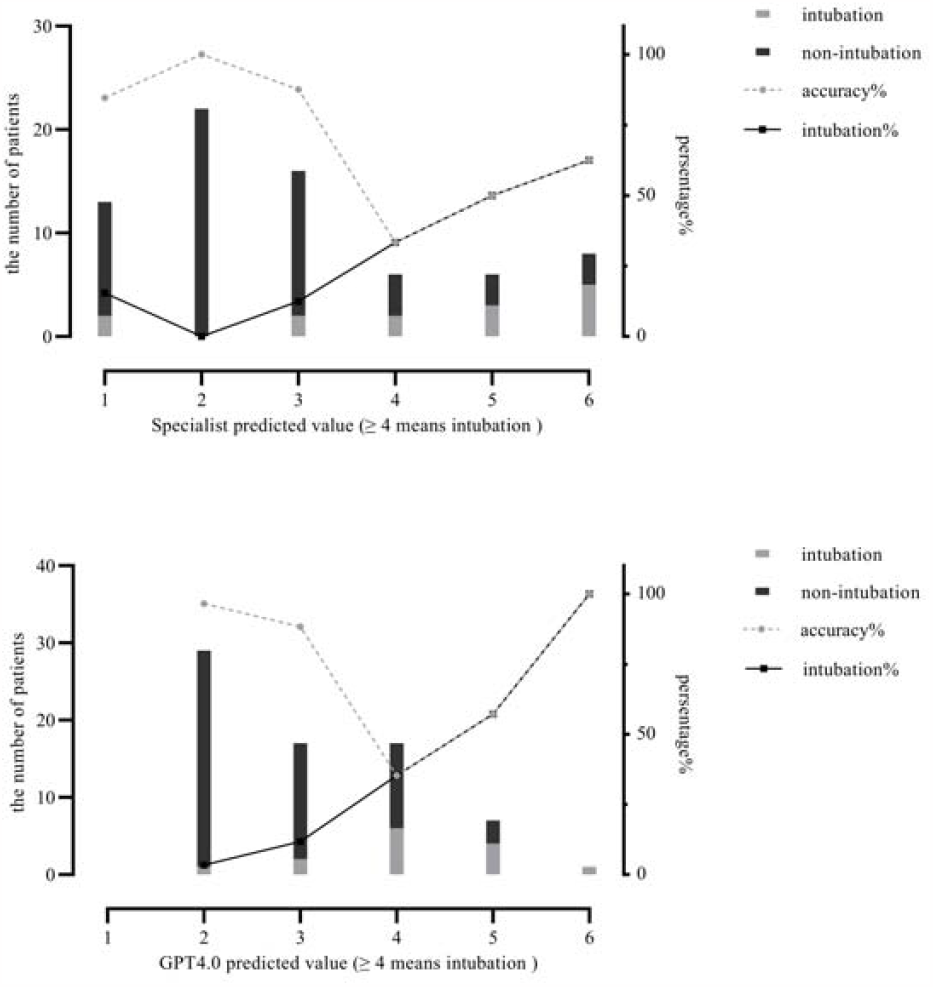
Distribution of accuracy in predicting endotracheal intubation within 48 hours between GPT 4.0 and specialist physicians.

The 6-hour ROX index, which is defined as (SpO_2_/FiO_2_)/respiratory rate[7], was calculated as a predictor for HFNC failure. We compared ROX prediction results as well as the results of the four questionnaires with the actual outcomes of 48-hour endotracheal intubation in patients. The receiver operating characteristic (ROC) curve was drawn and the area under the curve (AUC) was compared. The optimal diagnostic cutoff point was determined by the Youden index, and the overall accuracy, specificity, sensitivity, positive predictive value, and negative predictive value were calculated accordingly.

Patients was further divided into two groups based on the predicted values of specialist physicians and GPT 4.0 for prediction values ≥4 (high-risk group) and ≤3 (low-risk group), respectively. The 28-day cumulative endotracheal intubation curve and mortality curve were plotted and compared between the two groups of patients. The study flow was shown in Figure 1.

#### Box 1 The template of natural language questionnaire

The following is an illustration using data from a virtual patient. A female patient in her 60s was admitted to the hospital due to respiratory failure. The patient had not received mechanical ventilation treatment within the previous 24 hours. The patient had a history of cerebrovascular disease, and had no history of smoking.

The patient received high-flow oxygen therapy. At the beginning of high-flow oxygen therapy, the Glasgow Coma Scale was 8 points, the systolic blood pressure was 94 mmHg, the diastolic blood pressure was 46mmHg, the respiratory rate was 31 breaths per minute, the heart rate was 132 beats per minute, the pulse oxygen saturation was 88%, the oxygen flow rate was 40 L/min, the oxygen concentration was 54%, and blood gas analysis showed pH 7.5, pO_2_ 60mmHg, pCO_3_ 33 mmHg. The patient had received vasopressor medication.

After 6 hours of high-flow oxygen therapy, the patient’s systolic blood pressure was 82 mmHg, the diastolic blood pressure was 56 mmHg, the respiratory rate was 25 breaths per minute, the heart rate was 126 beats per minute, the pulse oxygen saturation was 91%, the oxygen flow rate was 40 L/min, the oxygen concentration was 55%, and blood gas analysis showed pH 7.46, pO_2_ 71 mmHg, pCO_2_ 3 mmHg. The patient had received vasopressor medication.

Please predict the risk of endotracheal intubation within 48 hours due to the failure of high-flow oxygen therapy according to the following options: 1. extremely unlikely to undergo endotracheal intubation; 2. unlikely to undergo endotracheal intubation; 3. Possible not to undergo endotracheal intubation; 4. possible to undergo endotracheal intubation; 5. likely to undergo endotracheal intubation; 6. extremely likely to undergo endotracheal intubation.

### Statistical Analysis

Sample size calculation: non-inferiority comparison using rate. According to the previous studies, the overall accuracy of 6-hour ROX index in predicting the 48-hour endotracheal intubation risk in patients is ∼0.8. The accuracy of non-specialist physicians’ prediction is estimated to be slightly lower, ∼0.75; The accuracy ChatGPT’s prediction is slightly higher and reaches ∼0.85. Therefore, Pt = 0.85, Pc = 0.75, δ = 0.1, and the ratio of sample sizes between the two groups is 1:1. Nc = Nt = 62. Considering ∼10% of patients being lost to follow up, 68 patients were planned to be included to create the questionnaires.

For normally distributed quantitative data, the arithmetic mean (standard deviation) is used, while for non-normally distributed data, the median (interquartile range) is used. Two independent sample t-tests and Mann-Whitney U tests are used for intergroup comparisons of continuous variables, and the chi-square test is used for rate comparisons. After drawing the ROC curve, the optimal diagnostic cutoff point is determined based on the maximum Youden index. The Log rank test is used to compare differences in 28-day mortality rates and to draw Kaplan-Meier survival curves. SPSS 26.0 is used for all data analysis, and GraphPad 8.0 is used for data visualization. P <0.05 indicates a statistically significant difference.

## Results

Among 71 patients, 14 patients (19.72%) required endotracheal intubation within 48 hours after HFNC therapy, 21 patients (29.58%) required endotracheal intubation within 28 days, and 16 patients (22.53%) died. There were no statistically significant differences between the intubation group and the non-intubation group in 48 hours after HFNC therapy in terms of baseline data, including age, gender, BMI, comorbidities, and other factors (all P>0.05). However, after 6 hours of HFNC oxygen therapy, the intubation group had significantly decreased pH, PaO_2_, and SpO_2_ (all P<0.05) compared to the non-intubation group (see Table 1).

The optimal diagnostic cut-off points for GPT-4.0 and specialist physicians were determined to be ≥4. The precision of GPT-4.0 was 76.1% [specificity=78.6% (95%CI=52.4-92.4%); sensitivity=75.4% (95%CI=62.9-84.8%)]. The positive predictive value was 40.7%, and the negative predictive value was 93.5%. The precision of specialist physicians was 80.3% [specificity=71.4% (95%CI=45.4-88.3%); sensitivity=82.5% (95%CI=70.6-90.2%)]. The positive predictive value was 50.0%, and the negative predictive value was 92.2%. The optimal diagnostic cut-off points for GPT-3.5 and non-specialist physicians were determined to be ≥5, with precisions of 73.2% and 64.8% respectively.

The area under the ROC (AUROC) of GPT-4.0 was 0.821 (95% CI=0.698-0.943), which was greater than, but not significantly (p>0.05) different from the AUROCs of GPT-3.5 [0.775 (95%CI=0.652-0.898)] and specialist physicians [0.782 (95%CI=0.619-0.945)], while was significantly higher than the AUROC of non-specialist physicians [0.662 (95%CI=0.518-0.805), P=0.011]. (Table 3 and Figure 3) Grouping the patients by GPT-4.0’s prediction value ≤3 (low-risk group, N=46) and ≥4 (high-risk group, N=25), the 28-day cumulative intubation rate (56.00% vs. 15.22%, P<0.001) and 28-day mortality (44.00% vs. 10.87%, P<0.001) were significantly higher in the high-risk group than in the low-risk group. There were statistically significant differences between the two groups of patients in terms of the parameters of heart rate, respiratory rate, pH, PaO2, SpO2, FiO2, and oxygen flow rate after 6 hours of HFNC therapy (all P<0.05). (Table 4 and Figure 4)

**Table 3.** Comparison of ROC area and accuracy for predicting endotracheal intubation.

|  | AUC<br>(95%<br>CI) | P | Cut-off | Sensitivity<br>(95% CI) | Specificity%<br>(95% CI) | positive<br>predictive<br>value | negative<br>predictive<br>value | Accuracy |
| --- | --- | --- | --- | --- | --- | --- | --- | --- |
| GPT4.0 | 0.821<br>(0.698-0.943) | - | ≥4 | 75.4%<br>(62.9-84.8%) | 78.6%<br>(52.4-92.4%) | 40.7% | 93.5% | 76.1% |
| GPT3.5 | 0.775<br>(0.652-0.898) | 0.484 | ≥5 | 71.9%<br>(59.2-81.9%) | 78.6%<br>(52.4-92.4%) | 40.7% | 93.2% | 73.2% |
| Sepecialist | 0.782<br>(0.619-0.945) | 0.475 | ≥4 | 82.5%<br>(70.6-90.2%) | 71.4%<br>(45.4-88.3%) | 50.0% | 92.2% | 80.3% |
| Non-Sepecialist | 0.662<br>(0.518-0.805) | 0.011 | ≥5 | 64.9%<br>(51.9-76.0%) | 71.4%<br>(45.4-88.3%) | 31.0% | 88.1% | 64.8% |
| ROX index | 0.746<br>(0.576-0.916) | 0.296 | ≤7.90 | 71.9%<br>(59.2-81.9%) | 78.6%<br>(52.4-92.4%) | 40.7% | 93.2% | 73.2% |

**Table 4.** Using GPT-4.0 to predict baseline data and prognosis of patients with and without endotracheal intubation.

|  | GPT4.0 ≤ 3<br>n=46 | GPT4.0 ≥ 4<br>n=25 | P |
| --- | --- | --- | --- |
| Age, mean (SD) | 68.26±15.35 | 69.24±15.58 | 0.800 |
| Male, n (%) | 30 (65.22%) | 15 (60.00%) | 0.663 |
| BMI, mean (SD) | 21.71±3.65 | 22.15±4.11 | 0.662 |
| Severe pneumonia, n (%) | 15 (32.61%) | 14 (56.00% ) | 0.055 |
| Type 1 respiratory failure, n (%) | 16 (34.78% ) | 8 (32.00%) | 0.813 |
| Sepsis, n (%) | 6 (13.04%) | 4 (16.00%) | 0.732 |
| Comorbidities |  |  |  |
| COPD, n (%) | 9 (19.57%) | 2 (8.00%) | 0.198 |
| Other chronic lung diseases, n (%) | 7 (15.22%) | 4 (16.00%) | 0.931 |
| Coronary heart disease, n (%) | 2 (4.34%) | 1 (4.00%) | 0.945 |
| Heart failure, n (%) | 4 (8.70%) | 2 (8.00%) | 0.920 |
| Chronic kidney disease, n (%) | 0 (0.00%) | 2 (8.00%) | 0.052 |
| Cerebrovascular disease, n (%) | 7 (15.21%) | 6 (24.00%) | 0.361 |
| Active tumor, n (%) | 3 (6.52%) | 4 (16.00%) | 0.201 |
| Smoking history, n (%) | 20 (43.48%) | 6 (24.00%) | 0.104 |
| Mechanical ventilation within the previous 48 hours, n (%) | 9 (19.57%) | 4 (16.00%) | 0.711 |
| When starting high flow oxygen therapy |  |  |  |
| GCS, median [Q1,Q3] | 15.0 [12.0, 15.0] | 12.0 [8.5, 14.5] | 0.008 |
| Heart rate, min<sup>-1</sup>, median [Q1,Q3] | 101.5 [92.5, 119.3] | 110.0 [83.0, 134.0] | 0.243 |
| Respiratory rate, min<sup>-1</sup>, mean (SD) | 25.76±7.90 | 29.48±8.81 | 0.073 |
| SBP, mmHg, mean (SD) | 121.46±24.12 | 125.04±22.09 | 0.540 |
| DBP, mmHg, mean (SD) | 73.48±16.56 | 70.04±12.50 | 0.368 |
| Receiving vasopressor, n (%) | 3 (6.52%) | 5 (20.00%) | 0.086 |
| pH, mean (SD) | 7.43±0.10 | 7.38±0.13 | 0.164 |
| PaO<sub>2</sub>, mmHg, median[Q1,Q3] | 60.0 [52.0, 72.3] | 51.0 [41.5, 63.5] | 0.014 |
| PaCO<sub>2</sub>, mmHg, median[Q1,Q3] | 38.0 [32.5, 42.5] | 32.0 [26.5, 47.0] | 0.297 |
| SpO<sub>2</sub>, mmHg, median[Q1,Q3] | 89.5 [84.8, 92.3] | 87.0 [82.5, 90.0] | 0.096 |
| FiO<sub>2</sub>, mmHg, median[Q1,Q3] | 47.5 [40.0, 55.0] | 40.0 [40.0, 47.5] | 0.042 |
| Flow, L/min, mean (SD) | 38.85±10.06 | 42.00±7.64 | 0.177 |
| High flow oxygen therapy at 6 hours |  |  |  |
| Heart rate, min<sup>-1</sup>, median[Q1,Q3] | 95.0 [86.8, 105.3] | 108.0 [89.0, 122.0] | 0.021 |
| Respiratory rate, min<sup>-1</sup>, mean (SD) | 20.72±5.06 | 26.76±8.14 | 0.002 |
| SBP, mmHg, mean (SD) | 123.37±15.65 | 122.20±23.80 | 0.826 |
| DBP, mmHg, mean (SD) | 73.63±10.92 | 70.16±11.70 | 0.217 |
| pH, mean (SD) | 7.43±0.70 | 7.36±0.15 | 0.025 |
| PaO<sub>2</sub>, mmHg, median[Q1,Q3] | 92.0 [73.5, 125.0] | 60.0 [53.0, 67.8] | <0.001 |
| PaCO<sub>2</sub>, mmHg, median[Q1,Q3] | 37.0 [33.0, 45.0] | 37.0 [29.3, 51.3] | 0.910 |
| SpO<sub>2</sub>, mmHg, median[Q1,Q3] | 97.5 [95.8, 99.0] | 92.0 [86.0, 93.0] | <0.001 |
| FiO<sub>2</sub>, mmHg, median[Q1,Q3] | 45.0 [40.0, 55.0] | 55.0 [50.0, 77.5] | 0.001 |
| Flow, L/min, mean (SD) | 38.59±8.48 | 43.40±5.90 | 0.014 |
| Outcomes |  |  |  |
| LOS in hospital, day, mean (SD) | 21.64±21.25 | 15.20±12.87 | 0.174 |
| Intubation rate in 48 hours, n (%) | 3 (6.52%) | 11 (44.00%) | <0.001 |
| Intubation rate in 28 days, n (%) | 7 (15.22%) | 14 (56.00%) | <0.001 |
| Mortality in 28 days, n (%) | 5 (10.87%) | 11 (44.00%) | <0.001 |

**Figure 3.**
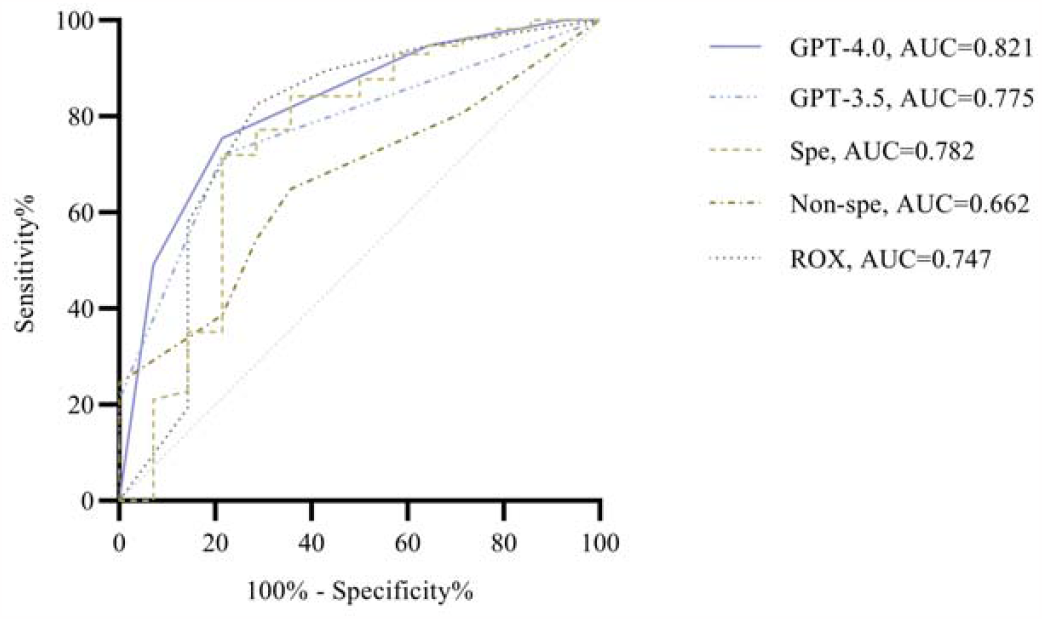
ROC curves of predicting endotracheal intubation within 48 hours for GPT and clinical physicians.

**Figure 4.**
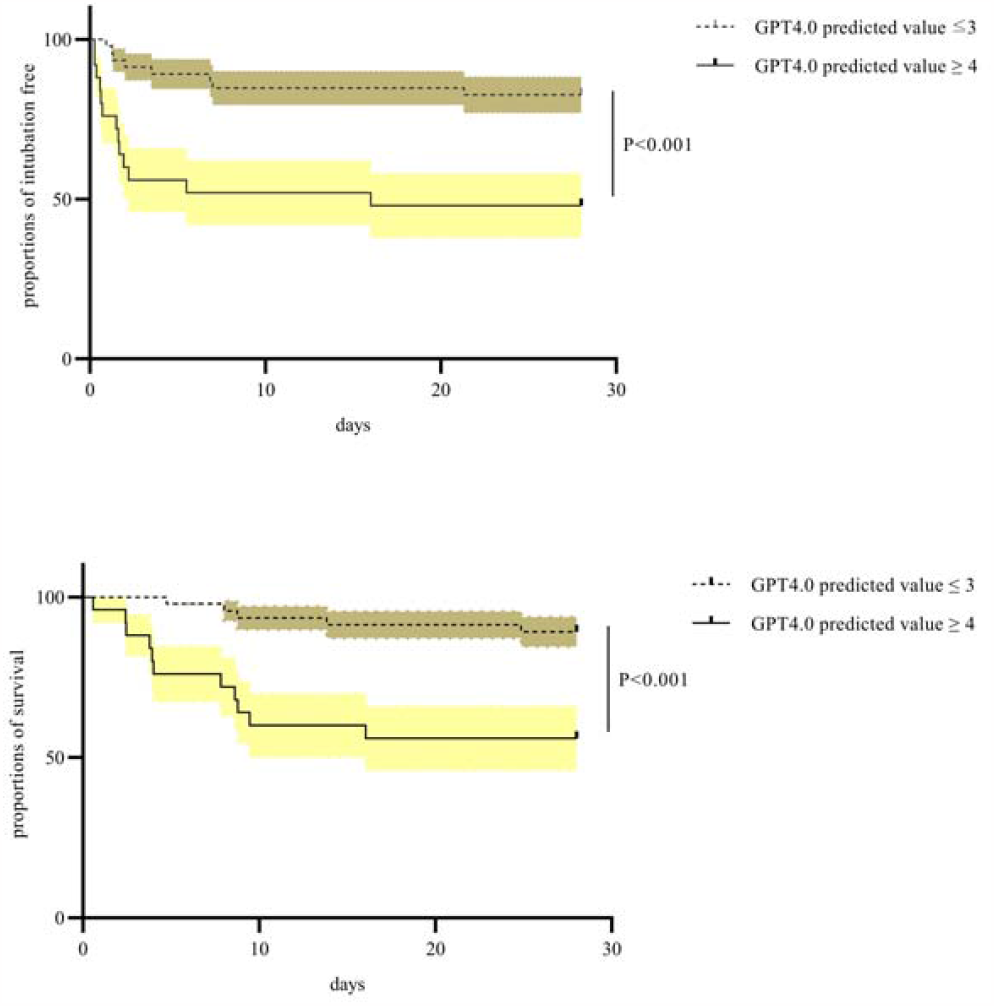
Cumulative endotracheal intubation curve and cumulative mortality curve of patients grouped by predicted values using GPT-4.0 at 28 days of treatment.

## Discussion

In this study, 71 patients with hypoxemia received HFNC oxygen therapy, and 14 of them underwent endotracheal intubation within 48 hours after HFNC therapy. We compared the predictive performance on 48-hour endotracheal intubation risk in patients for GPT-3.5, GPT-4.0, respiratory and critical care specialist physicians, and non-specialist physicians. The predictive AUROC of GPT-3.5, specialist physicians, and ROX index were all lower than 0.8.

For GPT-4.0, the optimal diagnostic cutoff point was determined to be ≥4 based on the maximum Youden index, which is in line with the natural language expression of the outcomes (3-Possible not to undergo endotracheal intubation; 4-possible to undergo endotracheal intubation). Its overall predictive accuracy had a good negative predictive value with an AUC of 0.821, and is significantly better than that of non-specialist physicians (0.662, P=0.011). Our results suggest that GPT-4.0 has clinical judgment experience that is at least better than that of non-specialist physicians on 48-hour endotracheal intubation risk in patients after HFNC therapy.

For both GPT-3.5 and non-specialist physicians, the optimal diagnostic cutoff point was determined to be ≥4, which should be ≥3 according to the natural language expression of the outcomes, indicating that GPT-3.5 and non-specialist physicians tend to overestimate the risk of tracheal intubation for patients due to the lack of experience. Besides, GPT-3.5 rarely gave answers of 1 and 6 in the questionnaire, indicating that it tends to give conservative answers whereas physicians tend to give a diverse range of answers based on their individual clinical judgment.

After grouping the patients according to the optimal diagnostic cut-off point of GPT-4.0, there were significant differences between the high-risk and low-risk patients in parameters such as heart rate, respiratory rate, pH, PaO_2_, SpO_2_, FiO_2_, and oxygen flow rate during the first 6 hours of high-flow oxygen therapy, indicating that GPT4.0 can effectively identify these clinical features to make accurate predictions.

It does not necessarily mean that the specialist physician’s prediction was wrong when their predictions do not match the actual clinical outcomes of patients within a short treatment period of 48 hours. This is because clinical practice is influenced by various factors, and the actual clinical physicians involved in the treatment may also make errors in judgments. However, when a study has adequate sample size, the accuracy of clinical physicians and GPT in predicting the endotracheal intubation risk can be compared based on the actual outcomes of patients.

When GPT predicted the endotracheal intubation risk for patients who had either a high or low intubation risk, its predictive accuracy was good. However, for patients whose intubation risk fell in the intermediate range, the overall accuracy of GPT’s prediction was only ∼50%. Specialist physicians and the ROX index also had poor predictive accuracy for these patients. While these are the patients who need early screening to avoid delayed intubation[9]. Since this study had a small sample size, subgroup analysis was not performed for these patients. Further analysis can be conducted in subsequent large-scale cohort studies.

The ROX index can be used to predict the failure of HFNC therapy. However, it only has a moderate level of predictive accuracy, and its diagnostic cutoff point lacks a unified standard as the ROX index only includes three parameters, namely the ratio of SpO_2_/FiO_2_ to respiratory rate[8, 10]. Incorporating more physiological parameters may improve the predictive accuracy[11, 12],therefore we expect to improve the predictive accuracy of endotracheal intubation risk by collecting more baseline data and physiological parameters of patients as well as using the algorithm model of ChatGPT. We argue that using GPT to predict the endotracheal intubation risk in patients receiving HFNC oxygen therapy is promising for clinical application[13]. GPT has a great potential to surpass the specialist physicians in terms of the clinical experience on judging endotracheal intubation risk after HFNC therapy[14]. Therefore, it can dynamically monitor patient data and reduce labor costs with its fast and convenient advantages[15].

We are concerned that GPT’s decision is based on accumulated data from actual clinical practice. However, if all clinical physicians rely on GPT to decide whether a patient should undergo endotracheal intubation in the future, the results will further strengthen GPT’s cognitive behavior. GPT would then become both the athlete and the referee. This would run counter to actual clinical needs. We cannot expect artificial intelligence to “grab its own hair to lift it off the ground.” Therefore, we need to prepare corresponding ethics for the clinical application of GPT[16, 17].

Limitations: 1. The answers of GPT are not entirely stable and can give different but similar answers for the same questionnaire. 2. This study is a multicenter prospective cohort study including only 71 patients, and a small number of specialist and non-specialist physicians answering the questionnaire. Therefore, with the progress of GPT model, a larger sample of patients should be included in clinical practice to further validate this conclusion.

## Conclusion

GPT-4.0 achieves an accuracy level comparable to specialist physicians in predicting the 48-hour endotracheal intubation risk in patients after HFNC therapy, based on patient baseline data and 6-hour parameters of receiving HFNC therapy. However, further large-sample studies are needed to inspect the reliability of using GPT-4.0 or more advanced version to support clinical decision.

## Data Availability

All data produced in the present work are contained in the manuscript.

## List of abbreviations

HFNC: high-flow nasal cannula
NIV: noninvasive ventilation
ICU: intensive care unit
BMI: body mass index
ABG: arterial blood gas
IQR: interquartile range
AUC: area under the receiver operating characteristic curve
ROX index: ratio of SpO_2_/FiO_2_ to respiratory rate

## Acknowledgments

Thanks to Shenyang RMS Medical Tech Company and Mr. Zhong Zhang for providing technical support.

## Authors’ contributions

Taotao Liu conceived the idea and led this study. Taotao Liu and Yaocong Duan interpreted the results and drafted the manuscript. Yanchun Li, Yingying Hu, Lingling Su and Aiping Zhang helped to interpret the results and drafted the manuscript. All authors read and approved the final manuscript.

## Consent for publication

Not applicable.

## Availability of data and material

All data generated or analyzed during this study are included in this article.

